# Epidemiological Parameters of SARS-CoV-2 in the UK during the 2023/2024 Winter: A Cohort Study

**DOI:** 10.1101/2024.07.22.24310801

**Authors:** Christopher E. Overton, Martyn Fyles, Jonathon Mellor, Robert S. Paton, Alexander M. Phillips, Alex Glaser, Andre Charlett, Thomas Ward

**Affiliations:** Department for Mathematical Sciences, University of Liverpool; Infectious Disease Modelling Team, All Hazards Intelligence, Data Analytics and Surveillance, UK Health Security Agency; Department for Electrical Engineering and Electronics, University of Liverpool; Statistics, Modelling, and Economics, Analytics & Data Science, Data Analytics and Surveillance, UK Health Security Agency

**Keywords:** COVID-19, sensitivity, incubation period, positivity, duration

## Abstract

Estimating epidemiological parameters is essential for informing an effective public health response during waves of infectious disease transmission. However, many parameters are challenging to estimate from real-world data, and rely on human challenge studies or mass community testing. During Winter 2023/2024, a community cohort study of SARS-CoV-2 was conducted across households in England and Scotland. From this survey, questionnaire data and follow-up testing protocols provided valuable data into multiple epidemiological parameters: namely, the duration of positivity, test sensitivity, and the incubation period. Here, Bayesian statistical modelling methods are developed and applied to estimate the underlying parameters. The duration of LFD positivity is found to increase with increasing age, with a mean of 8.55 days (95% CrI: 7.65 days, 9.44 days) in the youngest age group compared to 10.27 days (95% CrI: 9.85 days, 10.71 days) in the oldest age group. Similarly, test sensitivity, as a function of time since symptom onset, decays fastest in the youngest age group, reaching a minimum sensitivity of 0.26 (95% CrI: 0.16, 0.37) compared to 0.54 (95% CrI: 0.46, 0.6). Such patterns are expected since younger individuals experience less severe symptoms of COVID-19 and are likely to clear the virus faster. Combining the duration of positivity and test sensitivity, we estimate the probability of returning a positive test. Close to symptom onset date, this probability is approximately 95%. However, this rapidly drops off, dropping below 5% after 11.3 days (95% CrI: 9.7 days, 13 days) for the youngest age group and 16.2 days (95% CrI: 15.4 days, 17.1 days) for the oldest age group. For the incubation period, there is no clear pattern by age. Across all age groups, the mean incubation period is 2.52 days (95% CrI: 2.42 days, 2.62 days). This is shorter than the most recent estimates for Omicron BA.5, which is in line with earlier research that found replacing variants had shorter incubation periods.

## 1. Introduction

SARS-CoV-2, the virus that causes COVID-19, continues to cause resurgent global epidemics. The emergence of novel variants of SARS-CoV-2 has been driven by mutation from selective pressures and within-host factors. These mutations can lead to viral phenotypic changes that impact epidemiological parameters, such as the lower risk of mortality with Omicron relative to Delta [1–5], or shorter incubation periods of replacing variants [6–8]. To inform an effective public health response, understanding how these parameters change is vital for surveillance and policy.

During the height of the pandemic, mass testing data [9] and contact tracing data [10], alongside controlled studies, such as human challenge studies [11], were a valuable source of data on many epidemiological parameters, such as the incubation period and duration of positivity. Since April 2022, these surveillance efforts have been scaled down in the UK, with the Office for National Statistics (ONS) COVID-19 Infection Survey (CIS) concluding in March 2023. Similar reductions in surveillance data occurred globally, leading to uncertainty in these parameters. During Winter 23/24, the UK Health Security Agency (UKHSA) and ONS conducted a new community prevalence study to determine SARS-CoV-2 dynamics in England and Scotland. In this study, a randomly sampled cohort were tested using Lateral Flow Device (LFD) tests independently of symptom status to evaluate the trends of SARS-CoV-2 in the community [12]. However, as part of the study design, additional data were collected that provides valuable insights into multiple epidemiological parameters.

In this paper, data from this survey are used to estimate the duration of LFD positivity, LFD test sensitivity, and the current incubation period (the time from becoming infected to developing symptoms) of SARS-CoV-2. The most recent estimates of these parameters are limited to the pre-Alpha [13], Omicron BA.1 and BA.2 [14], and Omicron BA.5 [8,15] periods, respectively, reducing their utility in current public health policy. These parameters are essential in modelling/designing different interventions or public health messaging, as well as important parts of infectious disease surveillance tools. By providing estimates for these parameters during Winter 23/24, we gain insight into the current state of the virus.

## 2. Data

The UKHSA Winter Coronavirus Infection Survey (WCIS) builds on the success of the ONS CIS survey, and uses a subset of the same sample population. Around 150,000 individuals were invited to take part in the study, which ran from 13/11/2023 to 27/03/2024. The study design is described in the Supplementary Material.

Upon recruitment to the study, participants were provided with 14 SARS-CoV-2 LFD tests and were asked to complete a test every four weeks. Participants then had a 10-day window within which to complete those tests. Upon a participant testing positive, they were asked to complete a short questionnaire and to complete a follow-up testing protocol by continuing to test every other day until two consecutive negative results are observed. This repeat testing protocol, a modification from the design of the original ONS CIS, provides key data for estimating sensitivity and the duration of positivity.

A study participant is defined as an individual who returned at least one test result. There were 123,243 participants, of which 6,395 returned at least one positive test result. A total of 426,667 tests were performed as part of the main survey, of which 6,466 were positive. This averages 3.46 tests returned per participant that returned at least one test. This analysis is restricted to participants who returned at least one positive test result.

Unlike the original ONS CIS, participants were not compensated for their participation in the study. In addition, because of the re-use of the previous ONS CIS cohort, older individuals were over-represented in the sample. A demographic breakdown of the overall study participants is provided in the Supplementary Table 1. An age breakdown of study participants used in each model is provided in Table 1. Data inclusion criteria for each model are described in the corresponding methods sections.

**Table 1:**
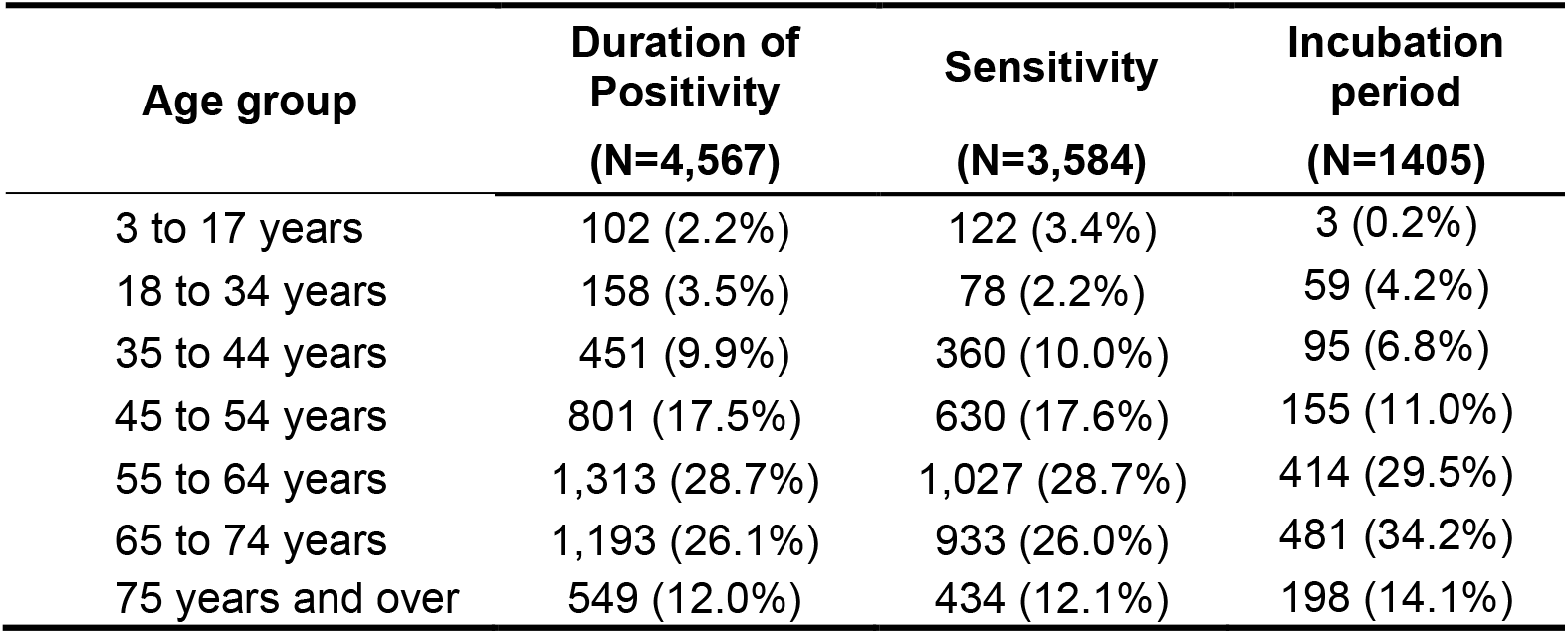
Age breakdown of the participants included in each model.

## 3. Methods

All methods used in this paper are Bayesian methods, implemented in the Stan [16] programming language, using CmdStanR [17] interfaced through R [18]. For each model, 4 chains were run generating 1000 samples each, with a warmup period of 1000 samples. Convergence of the models was assessed using the 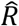 statistic [19], with a convergence threshold of 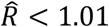.

### 3.1. Duration of LFD positivity

Understanding the duration an individual tests positive for is essential to understand the progression of a disease. For example, if isolation policies are based on individuals testing positive, it is important to know the duration of positivity, to determine whether this overlaps with the duration of infectiousness [20]. The duration of positivity is also vital to understand when calculating the incidence rate of new infections from a prevalence estimate.

The follow-up testing protocol provides interval-censored data on when individuals are no longer testing positive. Determining when an individual would start testing positive is challenging, since it will be left-censored at the date when they return their first positive test. To handle this, we instead calculate the duration of LFD positivity as a function of time since symptom onset date, which is reported in our data for 79% of individuals with at least one positive test. The symptom onset date is interval censored on the date that participants reported developing symptoms. We note, however, that only 8 positive tests occurred prior to the self-reported symptom onset of a case. This is expected since antigen levels only start to rise very close to symptom onset date [21].

For inclusion in the duration of positivity model data, data on an individual’s first positive test were linked to data from their follow-up testing. From this, the dates of the first positive test and last positive test were calculated. Data were cleaned by removing individuals with inconsistent testing dates. Of the 6395 individuals who submitted a positive test in the main survey, 4599 had a corresponding record in the repeat testing data. 32 records were removed due to inconsistent testing dates, leaving a final sample size of 4567 individuals. For the 8 individuals with positive tests prior to symptom onset date, we treat these as asymptomatic infections.

We consider two random variables, the time of symptom onset, *S*, and the time of last positive, *L*, where *L*>*S*. We are interested in the distribution of times between these two events, which we denote by the random variable τ ∈ ℝ_+_. Both types of data are considered interval censored:

- *S* ∈ [*s*_1,_ *s*_2_], where *s*_1_ = “Symptom onset date” and *s*_2_ = “Symptom onset date plus 24 hours”
- *L* ∈ [*l*_1,_ *l*_2]_, where *l*_1_ = “Last positive test date” and *l*_2_ = “First negative test date plus 24 hours”.

The 24 hours are added to each event since we only know the date and not the time of each event, so it could happen any time within that 24-hour window. Note that for *l*_2_, we only consider negative tests that occur after the last positive date, assuming that any earlier negatives are false negatives.

If the individual did not return any negative tests during their follow up testing, then *l*_2_ = ∞, i.e., the data are right-censored. If the individual does not have a symptom onset date, we treat the data as left-censored by the date of the first positive test, i.e., *s*_1_ = −∞ and *s*_2_ = “First positive test date plus 24 hours”. Based on these different censoring scenarios, there are four possible combinations that a data point can experience: (i) double-interval censoring, where the end points of each censoring interval are known; (ii) right-censoring, where the first event is interval censored and the second event is right-censored; (iii) left-censoring, where the first event is left-censored and the second event is interval censored; and (iv) left and right censoring, where the first event is left-censored and the second event is right censored. In each of these scenarios, we have a slightly different likelihood function. By considering distinct likelihood functions we make the model computationally feasible.

#### (i) Doubly-interval censored data

To model the duration of positivity, we wish to evaluate the following likelihood function

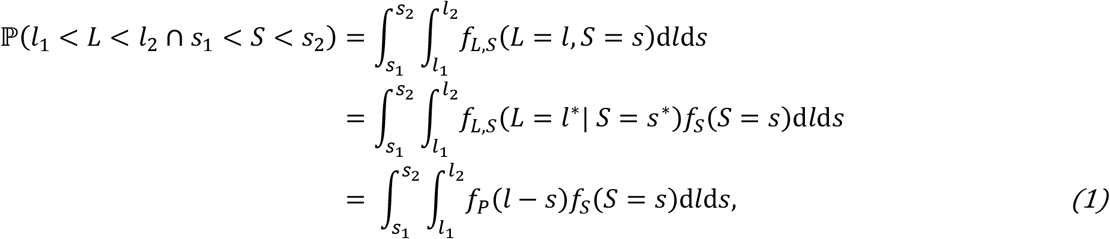

where *f*_*P*_ is the probability density function of the duration of positivity distribution. Implementing the likelihood in this form requires numerically evaluating a double integral, which is computationally expensive. Instead, we opt to use a latent variable approach [22–24], where we assume uniform prior distributions across the interval censored windows, i.e., 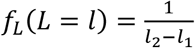 and 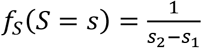, and sample the values of *S* and *L* from within their intervals by implementing:

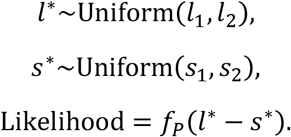

Here, since *l*^*^ follows a uniform distribution, adding the probability density function of *l*^*^ to the integrand in Equation (1) (which is implicitly done by the latent variable method) will not affect the results since it only adds a constant to the integrand.

#### (ii) Right-censored data

If our data are right-censored, the likelihood function is

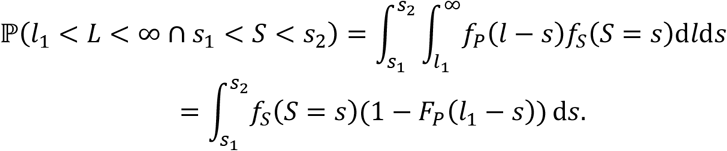

We again use a latent variable approach, but only a single latent variable is needed:

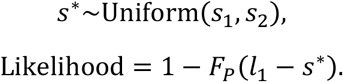

#### (iii) Left-censored data

Following similar logic to the right-censored model, if our data are left-censored, the likelihood function is

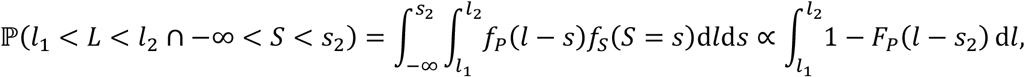

where an improper flat prior is implicitly used to model *f*_*S*_(*S* = *s*) = *c* ∀*s* ∈ [−∞, *s*_2_]. Using latent variables, we model this as

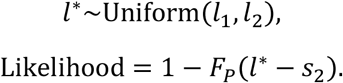

#### (iv) Left and right censored data

If the data are both left and right censored, we have

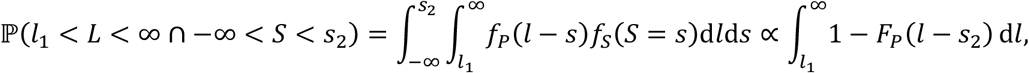

where an improper flat prior is implicitly used to model *f*_*S*_(*S* = *s*) = *c* ∀*s* ∈ [−∞, *s*_2_]. To solve this exactly would require numerically evaluating the integral or a latent variable approach with a uniform latent variable bounded between *l*_1_ to ∞. To avoid any issues with using such an improper prior, we can instead rewrite the model in terms of a single random variable

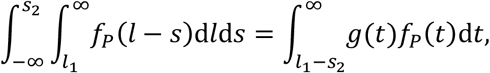

where *g*(*l* − *s*) is the probability density function of the random variable *L* − *S*. We can approximate this by assuming *g*(⋅) follows a uniform distribution, which gives

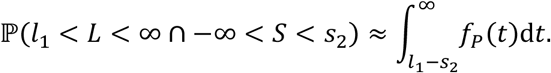

Therefore, the likelihood is given by

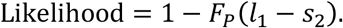

Under the assumptions of uniform prior distributions for *f*_*L*_(*L* = *l*) and *f*_*S*_(*S* = *s*), the random variable *L* − *S* follows a trapezoidal distribution [22,23]. Here, we approximate a trapezoidal distribution spanning from (*l*_1_ − *s*_2_, ∞) which with an improper uniform prior spanning (*l*_1_ − *s*_2_, ∞). This introduces some bias into the estimated variance [22], but has reduced computational cost and improved numerical stability relative to the latent variable approach.

Our model now consists of a loglikelihood that is significantly easier to evaluate, however there are many latent variables present in the model. As a result, while it is now possible to fit the model in a reasonable timeframe it cannot be considered fast. Many of these observations in the loglikelihood will have identical values of *s*_1,_ *s*_2_, *l*_1,_ *l*_2_. We now take advantage of this fact to develop an approximate model that vastly reduces the number of latent variables, and consequently provides a significant improvement to the model fitting speed.

In our loglikelihood we have terms of the form

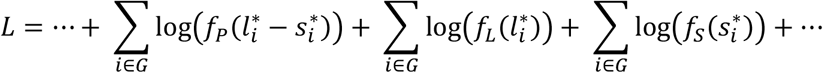

where *G* is a group of observations with identical values of *s*_1,_ *s*_2_, *l*_1,_ *l*_2_.

As an approximation, we will introduce random variables 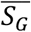 and 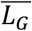, representing the means of the latent variables 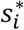 and 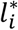 in group *G*. We will then approximate the loglikelihood as

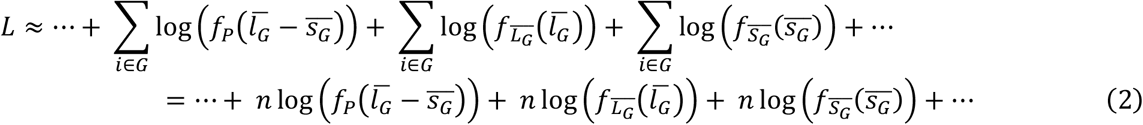

where *n* is the number of observations in group *G*. Since the latent variables 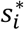 and 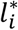 have standard uniform prior distributions, the sample mean of our latent variables, 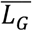 and 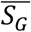, are given by Bates(*n*) distributions. Since the computational complexity of the Bates likelihood function increases with *n*, to improve numerical efficiency we will approximate the Bates(*n*) distributions using Beta distributions. For the Bates(*n*), we have

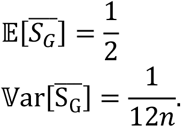

To approximate this, we construct a Beta distribution with the same mean and variance. For the Beta distribution, we have

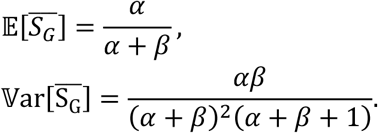

Equating the means gives

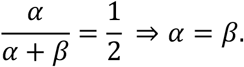

Equating the variances gives

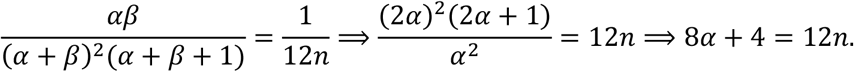

Therefore, we need 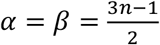. An alternative approximation would be to use a normal distribution, which is accurate for large *n* due to the central limit theorem. For *n* between 1 and 12, we compare the Beta and Normal approximation in Supplementary Figure 1. In general, both approximations perform well, but the Beta distribution works better for small values of *n*, in particular *n* = 1 where it exactly yields the Uniform(0,1) distribution, as desired. Therefore, we opt to use the Beta approximation throughout. In our preliminary analysis, using the grouped approximation (Equation (2)) was found to lead to indistinguishable results with a substantial reduction in computational time.

Regardless of the censoring scenarios, the likelihood of the model depends on the distribution of the positivity duration, either through the probability density function *f*_*P*_(⋅) or cumulative distribution function *F*_*P*_(⋅). The duration of positivity is assumed to follow a right-skewed distribution with a non-zero mode, since positivity cannot end instantaneously. Since viral load distributions vary with age [25–27], we assume this distribution can vary with age group. To model this distribution, we assume a lognormal distribution, parameterised by the log-mean, θ_1,*i*_, and log-standard-deviation, θ_2,*i*_, parameters, which represent the mean and standard deviation of the logarithm of the distribution, for age group *i*. The log-mean parameter is assumed to vary with age, following a hierarchical structure, and we assume the log-standard-deviation parameter is fixed for all age groups, i.e. θ_2,*i*_ = θ_2_ for all *i*. We model the parameters and latent variables using the following priors,

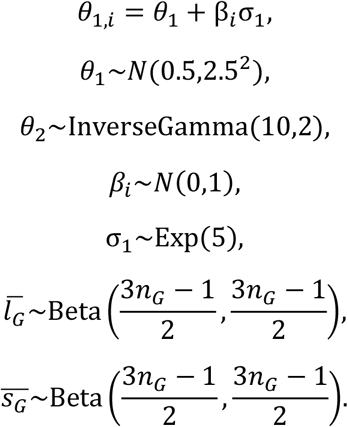

This hierarchical structure assumes that the log-mean parameter for each age group is centred around an average value, with perturbations specific to each age group. These perturbations are assumed to follow a normal distribution with mean of zero and variance of one. The sigma parameter controls the magnitude of the perturbations, i.e. larger values of σ_1_ allow individual age groups to have larger deviations from the population average. Therefore, where an individual age group has sparse data, the model will revert to the average value, and sufficient data is needed to justify large deviations from the average value. These age effects are additive on the log-mean parameter, which is roughly equivalent to a multiplicative effect on the linear scale (the effect is multiplicative on the median of the modelled distribution). For example, the influence of age on the median, relative to the population average, is a proportional change to the median duration of positivity. For the latent variable priors (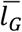 and 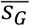), *n*_*G*_ is the number of observations in group *G*.

### 3.2. LFD test sensitivity

LFD tests are a powerful diagnostic tool due to a very short delay from testing to results. However, this comes at a compromise of reduced sensitivity to small viral loads relative to the gold-standard PCR tests. Therefore, when using LFD test results to monitor the epidemic, it is essential to quantify the corresponding test sensitivity.

Test sensitivity is typically given/interpreted as the probability of obtaining a positive test result, given that the tested individual is “currently infected”. There is, however, some ambiguity in how “currently infected” is defined. In an ideal world, we might define “currently infected” to be individuals who have not yet achieved viral clearance or who have a non-zero probability of transmission, though in practice it is impossible to determine whether individuals are in either of those states. Instead, surrogate definitions of “currently infected” are used. Most commonly, PCR positivity is used as a surrogate for “currently infected”, where an individual is defined as “currently infected” if there is a non-zero probability of returning a positive PCR result that is not a false positive [14,28]. We note that this definition may not correspond to viral clearance, since PCR tests can detect inactive viral fragments a significant length of time after the infection was cleared, nor does it necessarily correspond to actively infectious cases for the same reason.

Since no PCR tests were used in this study, it is impossible to estimate the probability of obtaining a positive LFD result given that an individual is PCR positive. Instead, our definition of “currently infected” must be individuals who have a non-zero probability of returning a positive LFD test result that is not a false positive. In other words, our definition of test positivity is the probability of obtaining an LFD positive result, given that an individual’s infection could be detected by an LFD test. As a result, if LFD sample positivity is adjusted to obtain the prevalence, the prevalence is defined as “the prevalence of LFD positive individuals”. We consider sensitivity as a function of time since symptom onset, as a proxy for time since infection. Therefore, this sensitivity tells us the probability of correctly returning a positive test for someone who is still testing positive a specific number of days after symptom onset. The period of the highest infectiousness is in the few days after symptom onset [21], which also corresponds to the period of high LFD test sensitivity, suggesting our definition of “currently infected” will capture the highly infectious individuals. We only consider sensitivity after symptom onset, since we are treating this as the time of earliest positivity.

For inclusion in the test sensitivity model, we filter the repeat testing follow-up to individuals who submitted an initial positive test in the main survey. For each individual, we observe a sequence of test results starting with a positive, e.g. “positive, positive, negative, positive, negative, negative” (Figure 1). The first test in this sequence will always be a positive result, as the individual would not be included in the repeat testing data had they not tested positive in the main survey. Therefore, we remove the first positive repeat testing result as it is not a random variable. It is assumed that once an individual begins to return consecutive negative tests they are no longer in the positive state, i.e. if the 6^th^ and 7^th^ tests are both negative, then the individual is no longer considered to be positive as of the 6^th^ test. Each testing sequence necessarily ends with a “positive, negative, negative” subsequence, which implies the test prior to the first observation of two consecutive negatives must be a positive test. Therefore, we also remove the last positive test as it is, by definition, a positive test result and cannot be treated as a random variable. Since we are interested in sensitivity conditional on still testing positive, all tests after the last positive test are also removed.

**Figure 1:**
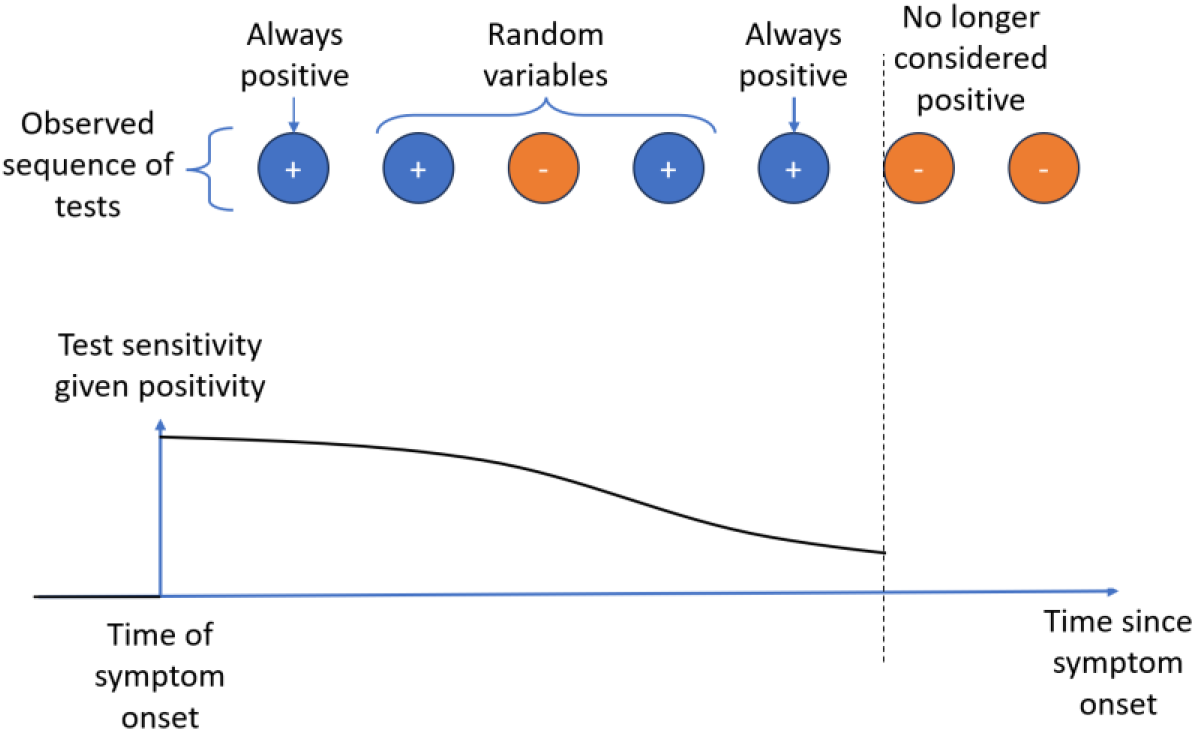
Schematic representation of the structure of the repeat testing data and model

This reduces the number of follow-up tests to 6758 tests. Individuals with unknown symptom onset date are removed, and individuals with negative time from symptom onset to test or symptom onset date 25 days or longer before testing date are also removed. This further reduces the number of positive tests to 6586, which forms our sample for the test sensitivity model. These tests were submitted by 3584 unique participants.

For each test included, we consider this data point as a pair of observations: the number of days from symptom onset, *d* ∈ ℤ_+_, drawn from a random variable *D*, and whether the test result, drawn from a random variable *r*, is positive (*r* = 1) or negative (*r* = 0). Since these data points are all conditional on the individual remaining in a positive state (*p* = 1), by aggregating the data by values of *D*, we can calculate the test sensitivity as a function of the time from symptom onset. Plotting the observed data (Supplementary Figure 2), we observe a sigmoidal shape to the data, with peak sensitivity at *D* = 0 and a plateau towards the end of the time period. This plateau occurs because we are conditional on individuals still testing positive, and restricting to a maximum of 24 days post symptom onset. If we increased the length of time since symptom onset, it is possible that a further decay in sensitivity would be observed. However, the probability of individuals still testing positive 25 days or longer after symptom onset is very low, which would lead to very few data points to inform the model. To capture the observed sigmoidal behaviour, we use a generalised logistic function

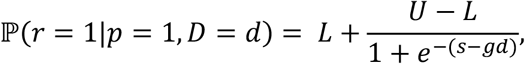

where *L* ∈ [0,1], *U* ∈ [0,1], *s* ∈ ℝ, and *g* ∈ ℝ_+_ are all parameters to be estimated. *L* and *U* represent the lower and upper bounds on the sensitivity, respectively. *s* is a parameter which shifts the curve, representing how soon sensitivity starts to decay. *g* is a rate parameter, which controls how quickly the sensitivity decays from the upper bound to the lower bound.

To fit the model, we aggregate the data such that for each value of *D*, we have the number of tests performed, *N*_tests_(*d*) ∈ ℤ_+_, and the number of positive tests that occurred, 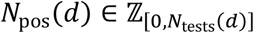, *d* days after symptom onset, in a population of cases that were still positive *d* days after symptom onset. We then assume that the observed number of positive tests is sampled from a binomial distribution with number of trials equal to total numbers of tests, and probability of success equal to ℙ(*r* = 1|*p* = 1, *D* = *d*), i.e.

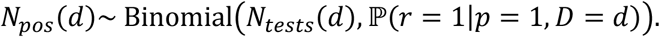

Test sensitivity will depend on the viral load of an individual at the time of their test. Since viral load trajectories will vary by age [25–27], test sensitivity is likely to vary with age [1]. To model this, we allow *L* and *g* to vary by age, and assume *U* and *s* are the same across all age groups, i.e.

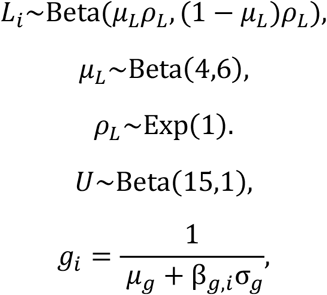

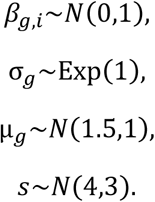

A version of the model allowing all 4 parameters to vary by age was developed, but the *U* and *s* parameters did not converge. The hierarchical structure for the rate parameter, *g*_*i*_, acts on the inverse as this aided convergence. This assumes that the inverse of the rate is affected by additive perturbations due to the different age groups, where *µ*_*g*_ is the population average inverse rate parameter and β_*g,i*_ are age group specific perturbations with magnitude controlled by σ_*g*_, which leads to non-linear perturbations on the scale of the rate. The hierarchical structure for the lower bound, *L*_*i*_, assumes that the lower bound for each age group is sampled from a Beta distribution with mean *µ*_*L*_, and the *ρ*_*L*_ parameter controls the variance of this Beta distribution. This ensures all lower bounds are bounded within the interval (0,1), and allows different age groups to have different lower bounds, but they are pulled towards the mean in the absence of data suggesting otherwise.

### 3.3. Probability of testing positive over time

Instead of considering test sensitivity, which is conditional on the individual being in the infected state, we may wish to calculate the probability that an individual returns a positive test result a certain number of days after their infection began. This differs from the test sensitivity definition, since this includes the probability that a negative test is returned because the individual has cleared the infection. This also differs from the duration of positivity, since that tells us the probability of whether an individual is still actively infected a certain number of days after infection began, rather than the probability that a single test will return a positive result. The probability of returning a positive a certain number of days after an infection is important, since it provides the relationship between epidemic incidence and sample positivity which then informs modelling of the epidemic trajectory.

As with the sensitivity and duration of positivity models, we will consider this as a function of time since symptom onset, rather than time since infection. Therefore, this tells us the probability that a test taken a certain number of days after the symptom onset date will return a true positive test result, using LFD test positivity as a proxy for infection. Previous studies [13,29–33] have shown that peak LFD sensitivity, which occurs around symptom onset, is comparable to peak PCR sensitivity, which is considered the gold standard at detecting infection. Therefore, this will accurately describe the probability of detecting an infection using LFD tests.

To calculate the probability of testing positive over time, we will use our results on test sensitivity and duration of positivity. Letting *S*(*d*) denote the probability of testing positive *d* days after symptom onset, we have

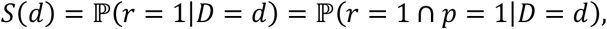

since an individual can only return a true positive test (*r* = 1) if it is still possible for them to test positive (*p* = 1). From this, we have

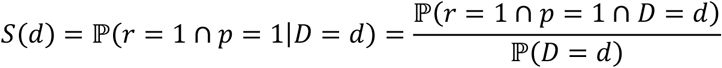

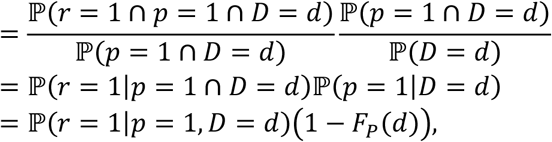

where ℙ(*r* = 1|*p* = 1, *D* = *d*) is the test sensitivity *d* days after symptom onset and 1 − *F*_*P*_(*d*) is the complimentary cumulative distribution function of the duration of positivity.

### 3.4. Incubation period

The incubation period describes the time between an individual becoming infected and developing symptoms. Information on time of infection is challenging to obtain. Commonly, contact tracing data has been used, where the date of contact between a primary and secondary case can be used as a date of exposure [34], however contact tracing for SARS-CoV-2 is no longer performed in the UK. In the absence of contact tracing data other approaches must be taken. For example, at the beginning of the COVID-19 pandemic, time spent in Wuhan could be used to provide an approximate exposure window [35–37]. In this study, we investigate the potential of asking individuals to identify their exposure date. For some individuals this may be relatively easy, for example if they have contact with a highly symptomatic individual or have infrequent contacts. For individuals with frequent potential infectious contacts, the data quality is likely to be lower. To improve the reliability of the reported exposure dates, we limit this analysis to individuals in either 1 or 2 person households. This restriction is made since in larger households it can be challenging to identify the index case, leading to incorrectly identified exposure dates. The secondary event, symptom onset date, is easy to measure, and we ask individuals to report when they first developed symptoms.

For inclusion in the data for the incubation period model, we first select individuals with an estimated exposure date, which reduced the sample size to 3949. We then filtered the data to individuals in households of size 1 or 2, reducing the sample size to 2767. Individuals with symptom onset date on the 15^th^ of any month were removed, since this was the default value if the question was not answered, reducing the sample size to 2089. Finally, individuals with inconsistent dates were removed, resulting in a final sample of 1405 individuals.

The incubation period describes the time between two epidemiological events, so we are interested in estimating the distribution of the possible time delays, since not all individuals will have the same value. In our data, the two events are time of exposure and time of symptom onset. Since not all infected individuals in the study will go on to develop symptoms, we only include an individual in our sample if they have developed symptoms. This introduces right truncation, whereby individuals with an exposure date within the study period but symptom onset date in the future were removed from our sample. This causes the observed time delay distribution to be biased towards shorter time delays. In this study, we are at the end of an epidemic wave, so the impact of the right-truncation will be minimal. In addition to the right truncation, our data are interval censored, since we only have an interval during which each event occurred, rather than the precise time. For both events, this interval censoring corresponded to a 24-hour window representing the day on which the event occurred. To correct for the interval censoring and right truncation, we use the Interval-censoring and right-truncation corrected approach (ICRTC) from [38]. Other potential approaches are described in [23], where this approach has been found to be the most accurate.

We consider two random variables, the time of exposure, *E*, and time of symptom onset, *S*, such that *E* < *S*. Our data takes the form of two intervals: the exposure window, *E* ∈ [*e*_1,_ *e*_2]_; and the symptom onset window, *S* ∈ [*s*_1,_ *s*_2]_, which each have a length of one day. The incubation period can be a described by a random variable *X* which we assume follows a positive-valued right-skewed distribution. We consider gamma, Weibull, and lognormal distributions, and assess goodness of fit through Pareto Smoothed Importance Sampling Leave-One-Out (PSIS-LOO) cross validation [39]. For each of these distributions, *X* is parameterised by two parameters, which we denote θ_1_ and θ_2_. To estimate these parameters, we consider the following interval-censoring and right-truncation corrected likelihood function

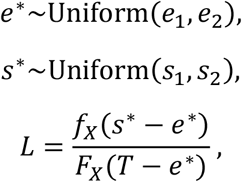

where *f*_*X*_ and *F*_*X*_ are the probability density function and cumulative distribution function, respectively, of the incubation period.

In addition to the right-truncation and interval-censoring corrections, in the data there is a second mode at zero, suggesting many respondents may have accidentally entered their symptom onset date as their exposure date. Therefore, we remove such individuals from the study. To account for this in the model, we additionally condition on the incubation period being larger than 1 day, which changes the likelihood function to

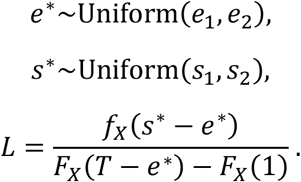

For each distribution, we parameterise the model such that θ_1_ represents the log of the mean. θ_2_ then represents the log of the standard deviation for the gamma and lognormal distributions, and the log of the shape parameter for the Weibull distribution.

Incubation periods potentially vary by age since the severity of symptoms is highly sensitive to age. To capture this, we consider a hierarchical version of the model, whereby θ_1_ and θ_2_ are age-specific parameters. We model this as

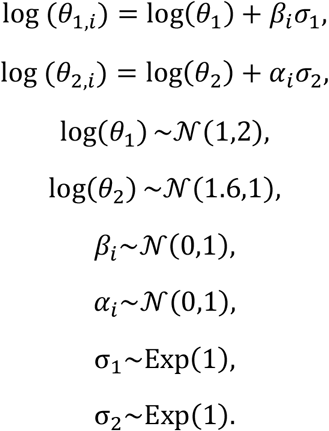

This assumes a similar hierarchical structure to the other models. The hierarchical assumptions are made on the logarithmic scale for the *θ*_1_ (mean) and *θ*_2_ parameters, which means that the influence of age is multiplicative on the scale of the model parameters. For example, the influence of age on the mean, relative to the population average, is a proportional change to the mean incubation period.

## 4. Results

### 4.1. Duration of positivity

For all age groups, the duration of positivity has very low early density (Figure 2) because very few individuals are expected to stop testing positive in fewer than 2 days. This is consistent with the raw data, where only 0.7% of doubly-interval censored data points had lower bounds on the duration of positivity less than 2 days. Looking across age groups, there was significant variation in the duration of positivity, though with high uncertainty in the youngest age groups (Figure 2, Table 2). The youngest age group, 3 to 17 years, has the shortest mean duration of positivity, estimated at 8.55 days (95% Credible Interval (CrI): 7.65 days, 9.44 days), and as age increases there is an increasing trend in the mean (with high uncertainty in some age groups), with the maximum mean duration of positivity in the oldest age group, 75 years and over, with an estimate of 10.27 days (95% CrI: 9.85 days, 10.71 days). In addition to the mean increasing with age, the upper percentiles of the duration of positivity also increases (Figure 3). For the 3 to 17 years age group, after 15.25 days (95% CrI: 13.63 days, 16.89 days), 95% of cases are no longer LFD positive. For the 75 years and over age group, this increases to 18.31 days (95% CrI: 17.51 days, 19.19 days).

**Table 2:** Mean, median, and 95^th^ percentile of the duration of positivity distribution. The given estimates are medians of the posterior distribution, and the numbers in brackets are 95% credible intervals.

| Age Group | Mean duration of positivity (days) | Median duration of positivity (days) | Duration of positivity 95<sup>th</sup> percentile (days) |
| --- | --- | --- | --- |
| 3 to 17 years | 8.55 (95% CrI: 7.65, 9.44) | 7.89 (95% CrI: 7.07, 8.71) | 15.25 (95% CrI: 13.63, 16.89) |
| 18 to 34 years | 8.82 (95% CrI: 8.16, 9.45) | 8.14 (95% CrI: 7.54, 8.73) | 15.74 (95% CrI: 14.53, 16.92) |
| 35 to 44 years | 9.24 (95% CrI: 8.82, 9.68) | 8.52 (95% CrI: 8.14, 8.93) | 16.47 (95% CrI: 15.67, 17.32) |
| 45 to 54 years | 8.93 (95% CrI: 8.60, 9.25) | 8.24 (95% CrI: 7.94, 8.53) | 15.92 (95% CrI: 15.29, 16.58) |
| 55 to 64 years | 9.43 (95% CrI: 9.18, 9.69) | 8.7 (95% CrI: 8.48, 8.94) | 16.81 (95% CrI: 16.28, 17.40) |
| 65 to 74 years | 9.66 (95% CrI: 9.40, 9.94) | 8.92 (95% CrI: 8.68, 9.17) | 17.24 (95% CrI: 16.68, 17.83) |
| 75 years and over | 10.27 (95% CrI: 9.85, 10.71) | 9.47 (95% CrI: 9.09, 9.88) | 18.31 (95% CrI: 17.51, 19.19) |

**Figure 2:**
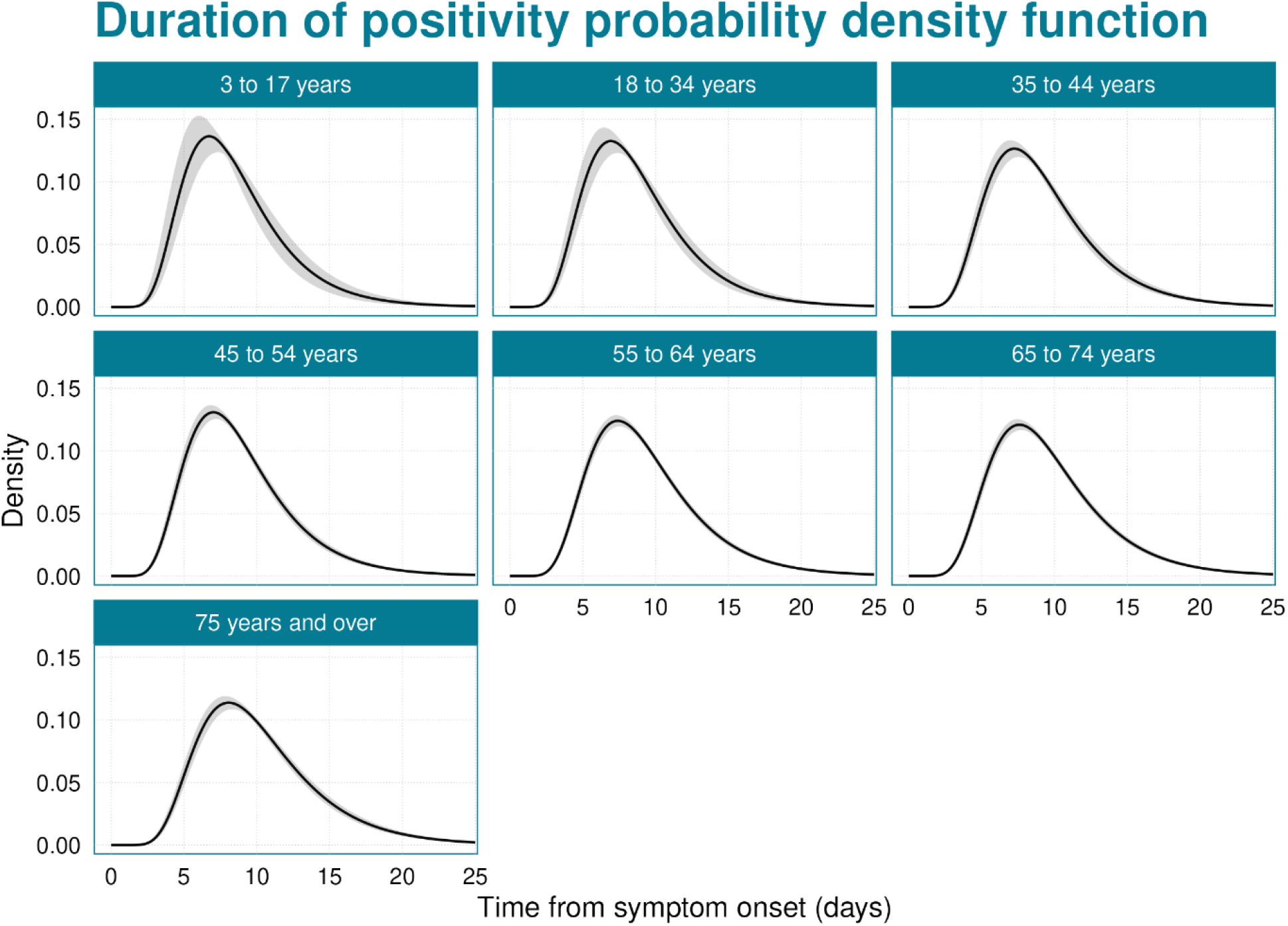
Probability density function of the duration of positivity. The black line shows the median of the posterior distribution, and the grey shaded region is the 95% credible interval.

**Figure 3:**
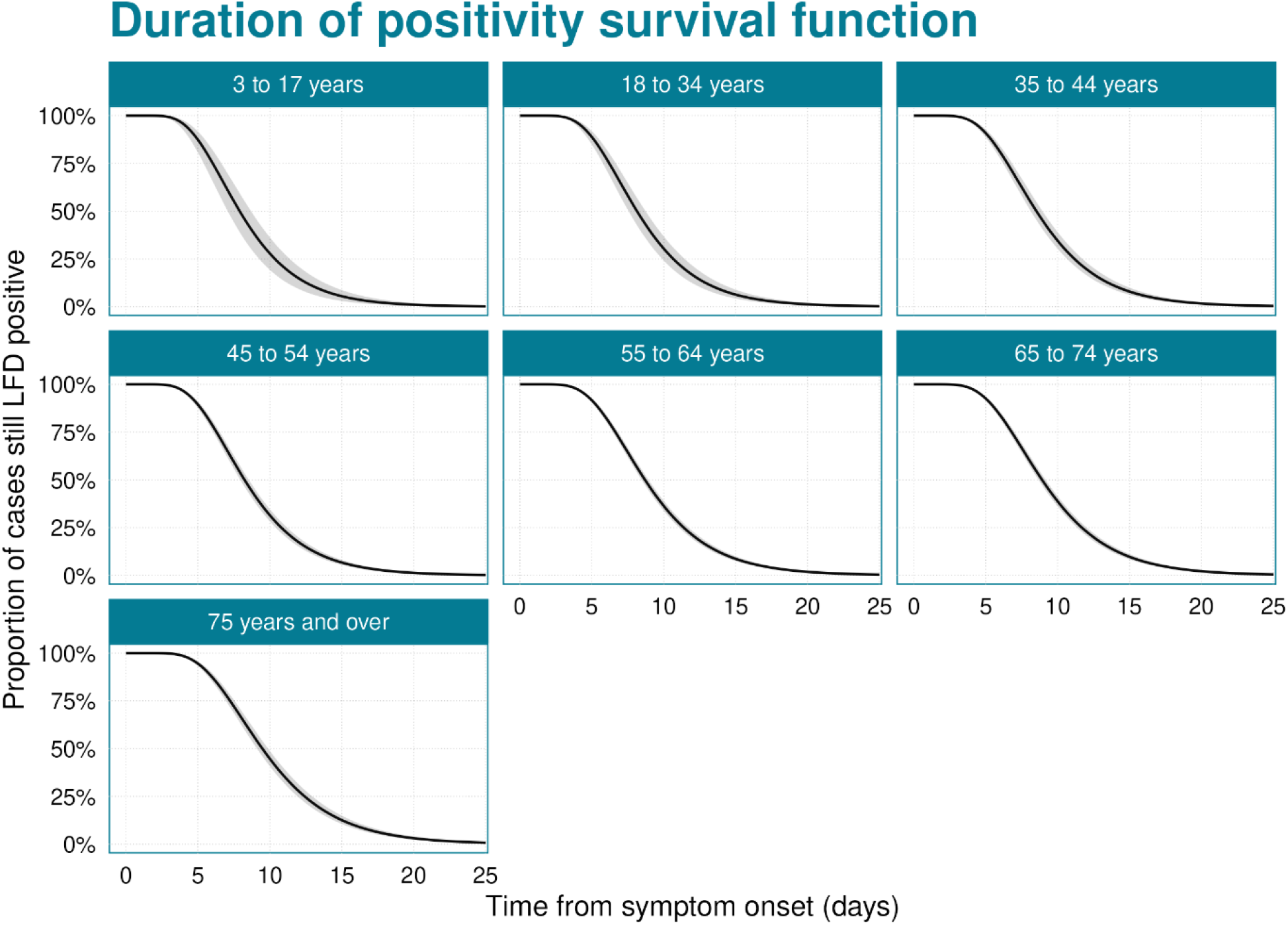
Probability that a case is still testing LFD positive. The black line shows the median of the posterior distribution, and the grey shaded region is the 95% credible interval.

No data are shown for goodness of fit comparisons due to the censored nature of the data, with each data point representing an interval of potential observations rather than a single observation, so empirical PDFs/CDFs cannot be calculated.

### 4.2. Sensitivity

Sensitivity is found to vary substantially with age (Figure 4, Table 3), with the minimum sensitivity increasing with increasing age. In the youngest age group, the minimum sensitivity is very low, plateauing at 0.26 (95% CrI: 0.16, 0.37). In the oldest age group, this increases to 0.54 (95% CrI: 0.46, 0.60). Peak sensitivity was assumed to be equal across all age groups, with a peak of 0.95 (95% CrI: 0.92, 0.98) at time of symptom onset. The rate at which sensitivity decays varied across age groups, with a pattern of decreasing decay rate with age. That is, not only does the youngest age group have the lowest lower bound, but it also reaches the minimum sensitivity the fastest.

**Table 3:** Estimated parameters of the sensitivity model. The given estimates are medians of the posterior distribution, and the numbers in brackets are 95% credible intervals.

| Age Group | Decay Rate | Lower bound | Upper bound | Shift |
| --- | --- | --- | --- | --- |
| 3 to 17 years | 1.05 (95% CrI: 0.79, 1.44) | 0.26 (95% CrI: 0.16, 0.37) | 0.95 (95% CrI: 0.92, 0.98) | 4.46 (95% CrI: 3.51, 5.66) |
| 18 to 34 years | 0.82 (95% CrI: 0.60, 1.12) | 0.42 (95% CrI: 0.30, 0.53) | 0.95 (95% CrI: 0.92, 0.98) | 4.46 (95% CrI: 3.51, 5.66) |
| 35 to 44 years | 0.73 (95% CrI: 0.57, 0.94) | 0.40 (95% CrI: 0.33, 0.47) | 0.95 (95% CrI: 0.92, 0.98) | 4.46 (95% CrI: 3.51, 5.66) |
| 45 to 54 years | 0.74 (95% CrI: 0.58, 0.96) | 0.34 (95% CrI: 0.27, 0.40) | 0.95 (95% CrI: 0.92, 0.98) | 4.46 (95% CrI: 3.51, 5.66) |
| 55 to 64 years | 0.78 (95% CrI: 0.61, 1.00) | 0.45 (95% CrI: 0.40, 0.49) | 0.95 (95% CrI: 0.92, 0.98) | 4.46 (95% CrI: 3.51, 5.66) |
| 65 to 74 years | 0.82 (95% CrI: 0.64, 1.04) | 0.51 (95% CrI: 0.46, 0.55) | 0.95 (95% CrI: 0.92, 0.98) | 4.46 (95% CrI: 3.51, 5.66) |
| 75 years and over | 0.67 (95% CrI: 0.52, 0.87) | 0.54 (95% CrI: 0.46, 0.60) | 0.95 (95% CrI: 0.92, 0.98) | 4.46 (95% CrI: 3.51, 5.66) |

**Figure 4:**
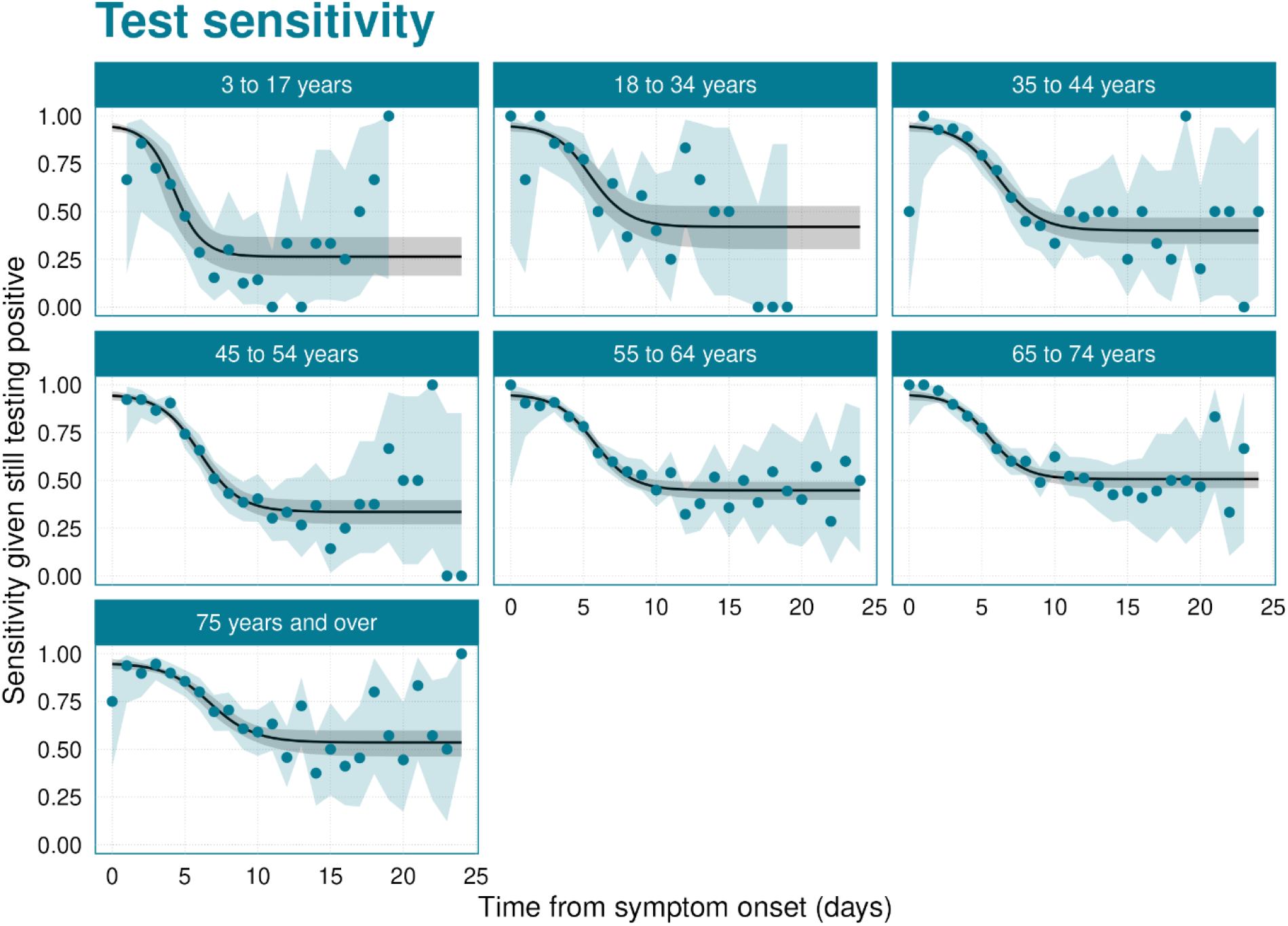
Test sensitivity given a case is still testing positive. The black line shows the median of the posterior distribution, and the grey shaded region is the 95% credible interval. The blue points are the raw data, and the blue shaded region indicates beta distributed 95% confidence intervals around the raw data.

### 4.3. Probability of testing positive

In the probability of testing positive, we observe a large increasing effect of age (Figure 5), since this combines the age signals observed in both the test sensitivity and duration of positivity parameters. The probability of testing positive rapidly drops off, as time from symptom onset increases, in the youngest age groups, with a considerably slower decline for the oldest age group.

**Figure 5:**
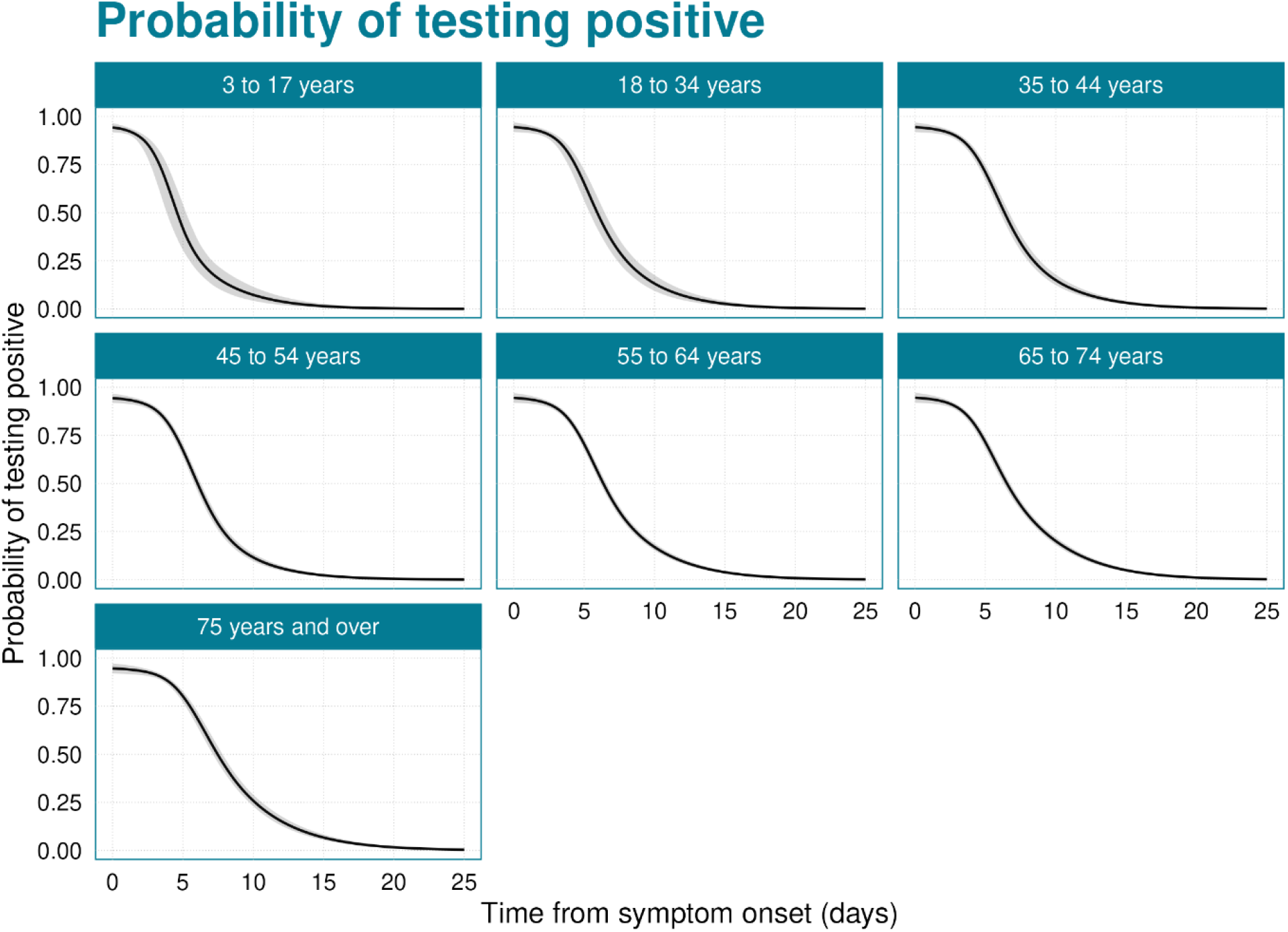
Probability that an individual returns a positive LFD test result. The black line shows the median of the posterior distribution, and the grey shaded region is the 95% credible interval.

At zero days from symptom onset, all age groups had 95% (95% CrI: 92%, 98%) probability of testing positive (Table 4). For the youngest age group, by 4.8 days (95% CrI: 4.0 days, 5.4 days) from symptom onset, this drops to 50% probability of testing positive, dropping below 5% probability after 11.3 days (95% CrI: 9.7 days, 13.0 days). For the oldest age group, these thresholds are increased to 7.6 days (95% CrI: 7.3 days, 8.0 days) and 16.2 days (95% CrI: 15.4 days, 17.1 days), respectively.

**Table 4:** Threshold values for time from symptom onset above which the probability of a test returning a negative result is above 50% and 95%. The given estimates are medians of the posterior distribution, and the numbers in brackets are 95% credible intervals.

| Age Group | Threshold where 50% of tests are negative (days) | Threshold where 95% of tests are negative (days) |
| --- | --- | --- |
| 3 to 17 years | 4.8 (95% CrI: 4.0, 5.4) | 11.3 (95% CrI: 9.7, 13.0) |
| 18 to 34 years | 6.1 (95% CrI: 5.5, 6.6) | 13.2 (95% CrI: 11.9, 14.4) |
| 35 to 44 years | 6.5 (95% CrI: 6.2, 6.8) | 13.7 (95% CrI: 12.8, 14.5) |
| 45 to 54 years | 6.2 (95% CrI: 6.0, 6.4) | 12.7 (95% CrI: 12.0, 13.4) |
| 55 to 64 years | 6.5 (95% CrI: 6.3, 6.7) | 14.3 (95% CrI: 13.8, 14.9) |
| 65 to 74 years | 6.7 (95% CrI: 6.4, 6.9) | 15.1 (95% CrI: 14.6, 15.7) |
| 75 years and over | 7.6 (95% CrI: 7.3, 8.0) | 16.2 (95% CrI: 15.4, 17.1) |

### 4.4. Incubation period

For the incubation period, there was no strong evidence of a difference between the model LOO scores estimated for each distribution (Supplementary Table 2). Weibull had the lowest estimated LOO, so all results are presented using a Weibull distribution, with gamma and lognormal distributions shown in Supplementary Table 3 and Supplementary Figures 3 and 4. There is no clear pattern in the incubation period with age (Figure 6, Table 5). Across all ages (Table 5), the mean incubation period was 2.52 days (95% CrI: 2.42 days, 2.62 days). The median incubation period was slightly shorter, at 2.24 days (95% CrI: 2.14 days, 2.35 days). The 95^th^ percentile of the incubation period, by which point we expect 95% of cases to have developed symptoms (Figure 7), was 5.53 days (95% CrI: 5.31 days, 5.79 days).

**Table 5:** Mean, median and 95^th^ percentile of the incubation period distribution. The given estimates are medians of the posterior distribution, and the numbers in brackets are 95% credible intervals.

| Age Group | Mean Incubation Period | Median Incubation Period | 95<sup>th</sup> Percentile of the Incubation Period |
| --- | --- | --- | --- |
| All ages | 2.52 (95% CrI: 2.42, 2.62) | 2.24 (95% CrI: 2.14, 2.35) | 5.53 (95% CrI: 5.31, 5.79) |
| 3 to 17 years | 2.55 (95% CrI: 2.25, 2.87) | 2.34 (95% CrI: 2.04, 2.65) | 5.25 (95% CrI: 4.44, 6.12) |
| 18 to 34 years | 2.57 (95% CrI: 2.39, 2.86) | 2.37 (95% CrI: 2.18, 2.65) | 5.31 (95% CrI: 4.83, 5.92) |
| 35 to 44 years | 2.54 (95% CrI: 2.35, 2.74) | 2.35 (95% CrI: 2.16, 2.56) | 5.12 (95% CrI: 4.65, 5.63) |
| 45 to 54 years | 2.50 (95% CrI: 2.31, 2.66) | 2.32 (95% CrI: 2.11, 2.49) | 5.03 (95% CrI: 4.60, 5.46) |
| 55 to 64 years | 2.50 (95% CrI: 2.36, 2.61) | 2.27 (95% CrI: 2.13, 2.39) | 5.26 (95% CrI: 4.96, 5.56) |
| 65 to 74 years | 2.62 (95% CrI: 2.50, 2.77) | 2.38 (95% CrI: 2.25, 2.53) | 5.54 (95% CrI: 5.25, 5.88) |
| 75 years and over | 2.53 (95% CrI: 2.36, 2.68) | 2.31 (95% CrI: 2.14, 2.47) | 5.25 (95% CrI: 4.89, 5.64) |

**Figure 6:**
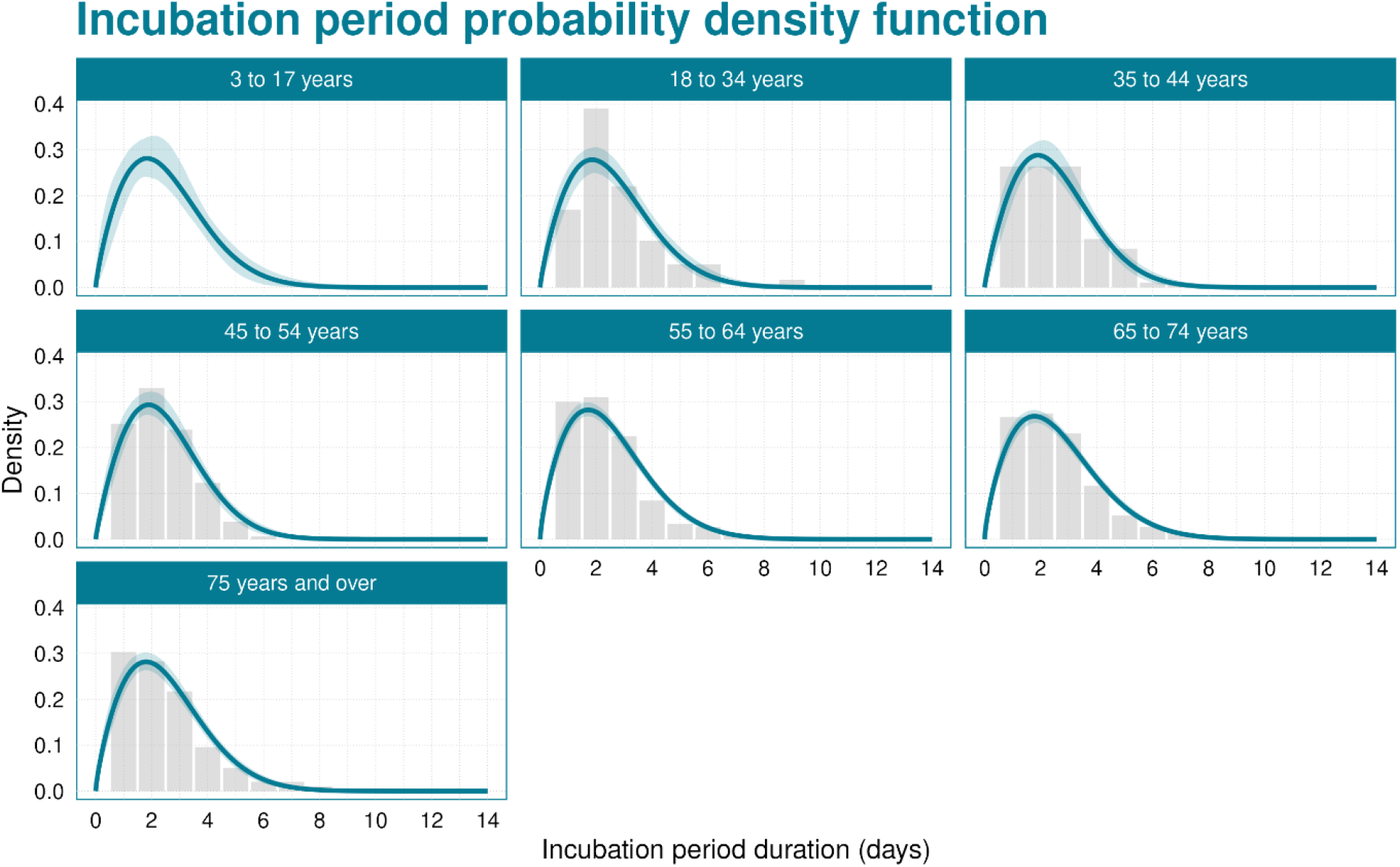
Probability density function of the incubation period. The blue line shows the median of the posterior distribution, and the blue shaded region is the 95% credible interval. The grey histogram shows the raw data (the 3 to 17 years age group data are masked due to low counts).

**Figure 7:**
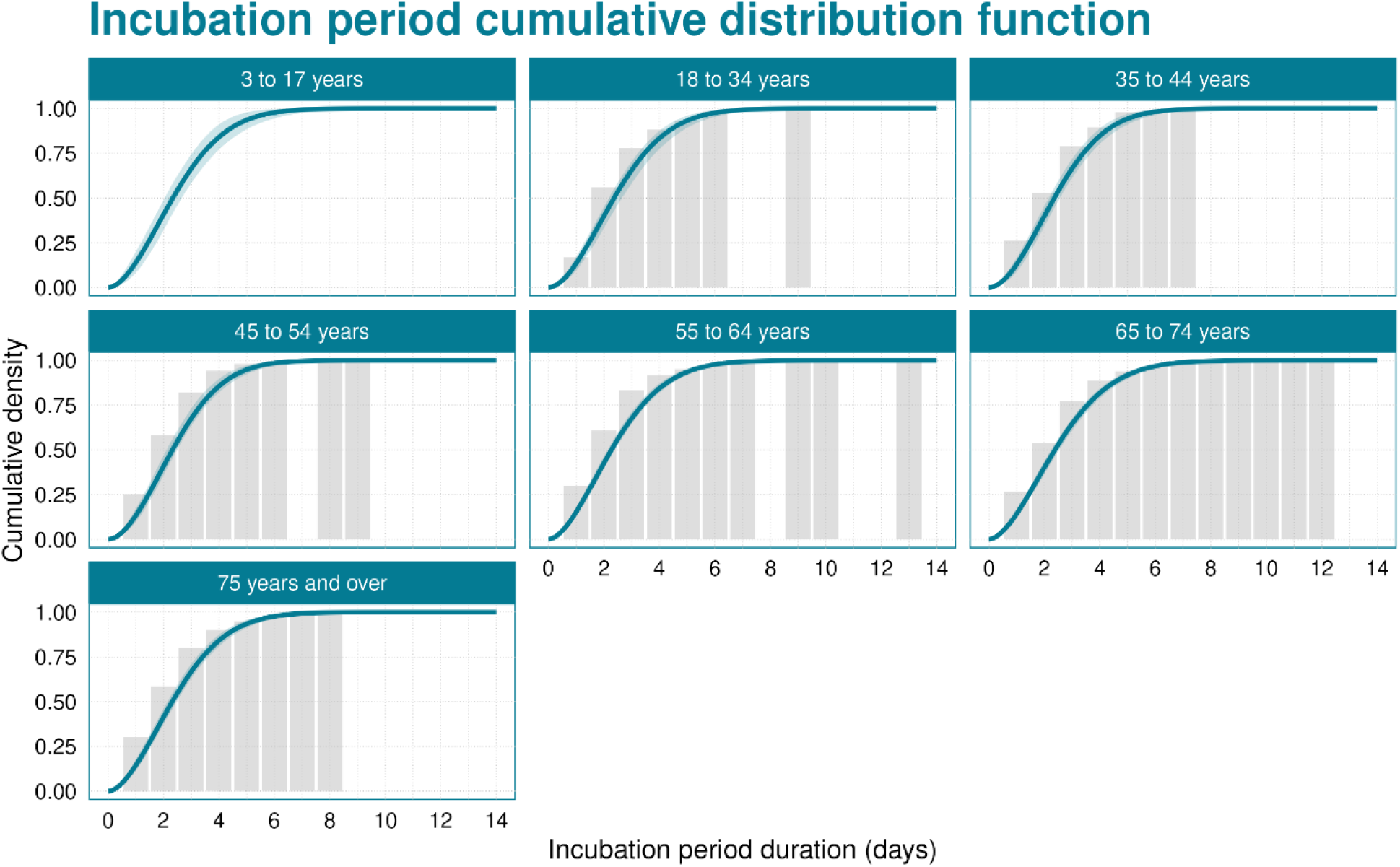
Cumulative distribution function of the incubation period. The blue line shows the median of the posterior distribution, and the blue shaded region is the 95% credible interval. The grey histogram shows the raw data (the 3 to 17 years age group data are masked due to low counts).

## 5. Discussion

The Winter Coronavirus Infection Survey in the UK has been a valuable source of information on the SARS-CoV-2 virus. From this survey, data were collected that can support the estimation of parameters that are of value to understanding the ongoing epidemiology of SARS-CoV-2. Given the complexities of the data and real-world biases, statistical modelling methods are needed to maximise the information extracted from the data. We have developed bespoke methods for estimating the duration of positivity and test sensitivity, and deployed the gold standard method for estimating the incubation period [23].

The duration of LFD positivity tells us how long after infection an individual remains able to test positive using LFD tests. This is important, as if an individual takes a test too long after infection, it may no longer be possible to test positive, and the infection will go undetected. Therefore, quantifying this distribution is vital when using testing data for infectious disease surveillance. Also, LFD positivity is closely linked to viral load in the individual [14]. The duration of positivity was shortest for the youngest age groups and increased with age. This finding is consistent with data on severity of infection, whereby older individuals are more likely to have severe disease [1,40,41]. Since severity is likely to correspond to higher viral load / slower viral clearance, this suggests that the duration of positivity should be shorter in younger individuals, which has been seen in earlier studies on the duration of viral shedding [25]. The duration of LFD positivity was found to have a mean of 8.55 days (95% CrI: 7.65 days, 9.44 days) days for the youngest age group, which increased with age to 10.27 days (95% CrI: 9.85 days, 10.71 days) in the oldest age group. Compared to previous estimates for pre-Alpha variants [13], this has reduced slightly from a median of 11 days (95% confidence interval: 10 days, 12 days) to a median of between 7.89 days (95% CrI: 7.07 days, 8.71 days) and 9.47 days (95% CrI: 9.09 days, 9.88 days), depending on the age of the individual. Such a reduction in the duration of positivity could be due to the reduced severity of Omicron sub-lineages [2–5] or higher levels of both infection-derived and vaccine-derived immunity. Based on the fitted lognormal distributions, 95% of cases are no longer LFD positive between 13.63 days and 19.19 days after symptom onset, depending on age. In our duration of positivity data, we do not have data on time of infection. Therefore, duration of positivity is considered as a function of time from symptom onset. However, in practice some individuals can test positive prior to symptom onset. In this study, such cases were very rare, suggesting that the probability of testing positive prior to symptom onset was very low, so this assumption will have a minimal influence on the results. The low probability of testing positive prior symptom onset is driven by antigen levels only growing shortly before symptom onset [21].

Test sensitivity for LFD tests has previously been shown to be lower than the gold-standard PCR tests [13,28,42]. However, despite the reduced sensitivity, due to the greatly improved time-to-result and reduced cost, LFD tests can be a powerful tool in the response to the COVID-19 pandemic [43]. The continued strength of LFDs relies on the test sensitivity remaining at similar levels, or increasing. Early findings for Omicron suggested increased sensitivity (73.0%) relative to Alpha and Delta variants (55.7% and 64.0%, respectively [14]). We estimated LFD test sensitivity as a function of time from symptom onset. Here, the sensitivity is relative to individuals who are still able to test LFD positive. It is possible that some individuals may never have high enough viral loads to test positive by LFD. However, previous estimates suggest that peak LFD sensitivity is comparable to peak PCR sensitivity [13]. LFD sensitivity at the peak was 95% (92%, 98%). This quickly decayed in all age groups, though the minimum sensitivity varied by age. In the youngest age group, the minimum sensitivity was lowest, at 0.26 (95% CrI: 0.16, 0.37). In the oldest age group, this increased to 0.54 (95% CrI: 0.46, 0.60). Not only did the youngest age groups have the lowest minimum sensitivity, but they also had the fastest rate of decay from maximum to minimum sensitivity. Higher sensitivity in older age groups supports the use of lateral flow tests in clinical settings [44], such as care homes, where the majority of patients are elderly. In this analysis, we considered sensitivity as a function of time since symptom onset. Often, sensitivity is reported as a single value [28,45]. From our temporal sensitivity, comparable single values can be generated by weighting the sensitivity distribution by the observed distribution of times from symptom onset for a given cohort. However, such figures are not reported in this analysis since they are highly dependent on the cohort distribution, which are not necessarily consistent between studies. Importantly, LFD test sensitivity in the few days proceeding symptom onset is very high, which overlaps when individuals are most infectious [21] and therefore enables efficient isolation of infectious individuals.

Based on the results for test sensitivity and duration of positivity, we calculated the probability of testing positive as a function of time since symptom onset. Whereas sensitivity gives the probability of testing positive given that someone is still infected, this instead gives the probability of testing positive given the number of days ago that the individual was infected. This provides the relationship between infection incidence and test positivity data, which is fundamental to accurate disease surveillance. We found that the effect of age was further amplified, with the youngest age group seeing rapid decline in the probability of testing positive, which slowed down as age increased. In the youngest age group, after 4.8 days (95% CrI: 4.0 days, 5.4 days), 50% of tests would no longer test positive, which increased to 7.6 days (95% CrI: 7.3 days, 8.0 days) in the oldest age group. After 11.3 days (95% CrI: 9.7 days, 13.0 days), fewer than 5% of tests in the youngest age group return a positive test result. This 5% threshold increases to 16.2 days (95% CrI: 15.4 days, 17.1 days) in the oldest age group. This shows that although LFD tests are reliable for detecting recent infection close to time of infection (using symptom onset as a proxy), their sensitivity rapidly declines so that they do not detect historic infections.

The incubation period describes the time between an individual becoming infected and eventually developing symptoms. This is important in the design of isolation strategies, for example, where we need to know how quickly people will develop symptoms after infection. At the start of the SARS-CoV-2 pandemic, incubation period estimates ranged from 4.84 days to 6.4 days [35–37,46]. The mean incubation period in this study was estimated at 2.52 days (95% CrI: 2.42 days, 2.62 days). This is shorter than estimates of 2.6 days to 3.8 days for Omicron BA.5 [8,15], which is the most recent variant with estimates in the literature. However, the incubation period was on a decreasing trajectory since wild type [6,8], and with countless variants emerging since Omicron BA.4/5, it may be possible for the incubation period to have decreased further. For example, influenza incubation periods are even shorter at 1.71 days [47]. With this incubation period, 95% of individuals have developed symptoms by 5.53 days (95% CrI: 5.31 days, 5.79 days). We found no patterns of changing incubation periods with age, which is consistent with some previous studies [6], though others have identified a weak relationship with age [48]. With the relatively small sample size, such a signal would be hard to detect. The incubation period data relies on individuals estimating their time of infection, which may introduce a bias. For some individuals, this may be accurate, such as those who infrequently leave the house. For individuals in large households, these data are likely to be inaccurate due to large number of potential infectors. To mitigate for this, we restricted the analysis to 1 and 2 person households. However, in 2 person households, it is possible that pre-symptomatic transmission can occur, which might bias the estimated exposure dates, leading to an underestimate of the incubation period.

## 6. Conclusion

The Winter Coronavirus Infection Study, although designed to improve our understanding of SARS-CoV-2 prevalence in the community, has been a valuable source of data on key epidemiological parameters. The study design used will be powerful in future community surveillance studies for allowing continued estimation of these parameters. Through this study, we have identified large changes in the epidemiology. Firstly, the duration of LFD positivity for currently circulating variants has reduced relative to pre-Omicron variants. Secondly, LFD test sensitivity remains high, particularly shortly after symptom onset, when individuals are likely to be the most infectious. Finally, the incubation period has been observed to have declined relative to earlier variants. Understanding the current values of these parameters is essential for designing policy and interventions, as well as for accurately converting test positivity data to estimates of infection incidence and prevalence, which are vital for assessing real-time infection risk in the community.

## Data Availability

Stan code for all models and trace files from the MCMC samplers are available at https://github.com/OvertonC/winter_covid_infection_study-parameters.
UKHSA operates a robust governance process for applying to access protected data that considers: 
-the benefits and risks of how the data will be used
-compliance with policy, regulatory and ethical obligations 
-data minimisation 
-how the confidentiality, integrity, and availability will be maintained 
-retention, archival, and disposal requirements 
-best practice for protecting data, including the application of privacy by design and by default, emerging privacy conserving technologies and contractual controls 
Access to protected data is always strictly controlled using legally binding data sharing contracts. 
UKHSA welcomes data applications from organisations looking to use protected data for public health purposes. 
To request an application pack or discuss a request for UKHSA data you would like to submit, contact.

https://github.com/OvertonC/winter_covid_infection_study-parameters

## Ethics Approval

The study received ethical approval from the National Statistician’s Data Ethics Advisory Committee.

## Conflict of Interest

The authors have declared that no competing interests exist. There was no financial support for this work, completed as part of the authors’ employment.

## Acknowledgements

The authors would like to thank the UKHSA Surveillance and Immunity team and ONS WCIS analysis team for their contributions.

## Data Availability Statement

Stan code for all models and trace files from the MCMC samplers are available at https://github.com/OvertonC/winter_covid_infection_study-parameters.

UKHSA operates a robust governance process for applying to access protected data that considers:

- the benefits and risks of how the data will be used
- compliance with policy, regulatory and ethical obligations
- data minimisation
- how the confidentiality, integrity, and availability will be maintained
- retention, archival, and disposal requirements
- best practice for protecting data, including the application of ‘privacy by design and by default’, emerging privacy conserving technologies and contractual controls

Access to protected data is always strictly controlled using legally binding data sharing contracts. UKHSA welcomes data applications from organisations looking to use protected data for public health purposes.

To request an application pack or discuss a request for UKHSA data you would like to submit, contact.

## Notes

### Competing Interest Statement

The authors have declared no competing interest.

### Author Declarations

National Statisticians Data Ethics Advisory Committee of UK Statistics Authority gave ethical approval for this work

