## Supplementary Material for "Epidemiological Parameters of SARS-CoV-2 in the UK during the 2023/2024 Winter: A Cohort Study"

**Winter COVID-19 Infection Study Design**

Cohort

The cohort sampled for this study was selected from previous participants of the COVID-19 Infection Study (CIS) (https://www.ons.gov.uk/peoplepopulationandcommunity/healthandsocialcare/conditionsanddiseases/bulletins/coronaviruscovid19infectionsurveypilot/latest), a household study. Participants were asked if they would consent to being contacted by UKHSA for future studies – those who selected yes were asked to enrol. Individuals eligible for the study lived in private households and were 3 years and older living in England or Scotland. Participants are asked a range of information about themselves and the results of SARS-CoV-2 diagnostic tests. The study was live from November 2023 to March 2024. Overall, there were 123,243 participants and 426,667 responses. Further information about how this data was collected can be found in the ONS quality and methodology information page for the study (https://www.ons.gov.uk/peoplepopulationandcommunity/healthandsocialcare/conditionsanddiseases/methodologies/wintercoronaviruscovid19infectionstudyqmi). Information for the participants, including how to complete the questionnaire and conduct the tests are found on the ONS website (https://www.ons.gov.uk/surveys/informationforhouseholdsandindividuals/householdandindividualsurveys/wintercoronaviruscovid19infectionstudy). The participant breakdowns by reference group are given in Supplementary Table 1.

Survey Design

The study had three core surveys, the first "participant" survey collected key identifying information used to link data, such as names, addresses and demographics. The second "main" survey asked further questions relating to individual characteristics (such as job, household size), current and new symptoms, key survey dates (including window start date, LFD taken date, symptom onset date) and lateral flow device (LFD) test results. The third "follow up" survey was given to those who reported a positive test in the "main" survey. The "follow up" survey asked participants to test on alternating days following their first positive test.
For each survey participants responded via an online survey portal which guided them through each question. Participants were not offered financial incentives for the study but were given the lateral flow devices as part of the operations.

Participants were grouped into waves, and testing windows. Each participant received 14 tests and the start of the study and responded once to the main survey per wave, a total of 4 times over the course of the study. Each wave ran for approximately 4 weeks, with wave 1 running from 14 November to 14 December 2023, wave 2 from 12 December 2023 to 11 January 2024, wave 3 from 9 January to 8 February 2024 and wave 4 from 6 February to 7 March 2024. To allow for flexible timing in testing and response participants were given a window of 7 days to report their test. Participants were prompted at the start of their testing window and again during their window to encourage response. Tests taken up to two days before the start of the testing window were allowed to be reported in the main survey.

Data Processing

Data were cleaned and duplicates dropped such that there was one record per participant per wave. Records were removed where linkage information was unavailable, or demographics information not given. This accounted for <1% of the participants and records. Samples were removed where the individual did complete the main survey but did not take an LFD test.
Where a participant did not know their symptom onset date they were prompted to give a placeholder date of the 15th of the most recent month, for both the follow up and main survey. Therefore, values for symptom onset on these dates were assumed as missing. Demographic and participant characteristic information was captured in the first main survey response and not again unless there was a change in circumstance to avoid lengthy surveys. These demographic data were therefore filled forward. Participants who withdrew consent part way through or after the study were excluded from the analysis.

***Supplementary Table 1***. *The count (n) and proportion (%) of participants in each strata. The proportions are calculated as the count within a strata divided by the count within a group.*

| group | strata | n (participants) | % (participants) | n (responses) | % (responses) | % (population) |
|---|---|---|---|---|---|---|
| all | | 123243 | | 426667 | | |
| age | 3 to 17 | 3886 | 3.15 | 11083 | 2.6 | 20.55 |
| age | 18 to 34 | 4220 | 3.42 | 11426 | 2.68 | 21.37 |
| age | 35 to 44 | 9959 | 8.08 | 29803 | 6.99 | 13.49 |
| age | 45 to 54 | 18448 | 14.97 | 61061 | 14.31 | 12.72 |
| age | 55 to 64 | 30387 | 24.66 | 108950 | 25.54 | 13.09 |
| age | 65 to 74 | 36813 | 29.87 | 133983 | 31.4 | 9.77 |
| age | 75 and over | 19530 | 15.85 | 70361 | 16.49 | 9 |
| sex | Female | 70189 | 56.95 | 241721 | 56.65 | 50.99 |
| sex | Male | 53054 | 43.05 | 184946 | 43.35 | 49.01 |
| location | North West | 13812 | 11.21 | 47568 | 11.15 | 12 |
| location | East Midlands | 8977 | 7.28 | 31266 | 7.33 | 7.86 |
| location | South East | 18782 | 15.24 | 65579 | 15.37 | 14.94 |
| location | London | 18118 | 14.7 | 61023 | 14.3 | 14.31 |
| location | South West | 12732 | 10.33 | 44551 | 10.44 | 9.19 |
| location | Scotland | 11469 | 9.31 | 39968 | 9.37 | 8.72 |
| location | East of England | 13969 | 11.33 | 48642 | 11.4 | 10.25 |
| location | Yorkshire and The Humber | 10634 | 8.63 | 36931 | 8.66 | 8.84 |
| location | West Midlands | 9722 | 7.89 | 33590 | 7.87 | 9.63 |
| location | North East | 5028 | 4.08 | 17549 | 4.11 | 4.27 |

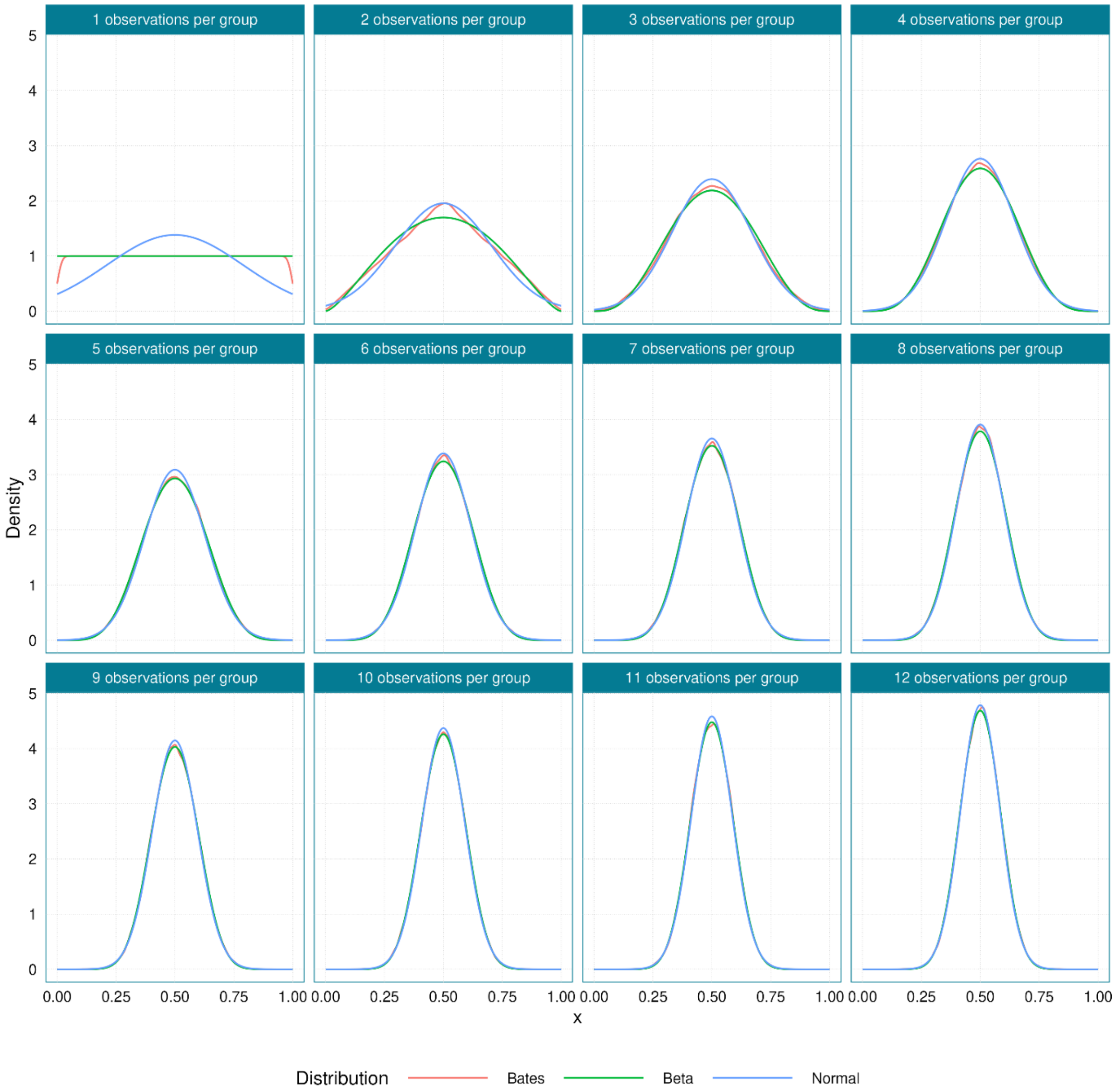


***Supplementary Figure 1:*** *Comparing the Beta and Normal distribution approximations to the Bates distribution, for Bates(1) to Bates(12) distributions. The Bates distribution densities are simulated. For Bates(1), the decline at 0 and 1 are numerical errors from approximating the density of a Uniform(0,1) distribution – the true density should be straight line from 0 to 1.*

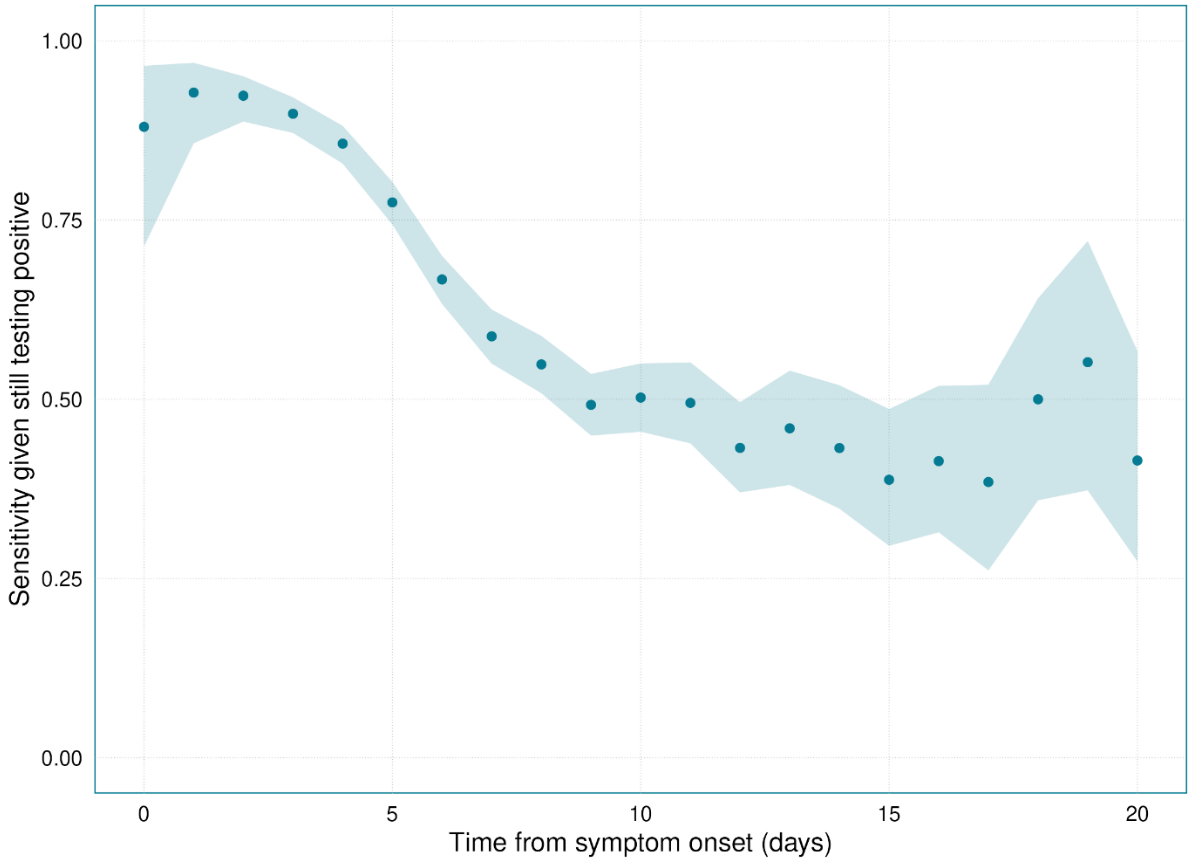


***Supplementary Figure 2:*** *Sensitivity data with beta distribution uncertainty, aggregated across all age groups. The dots are the raw data and the blue shaded region indicates beta distributed 95% confidence intervals around the raw data.*

***Supplementary Table 2:*** *Loo estimates for the three different distributions used to model the incubation period.*

| **Model** | **LOO – Estimate** | **LOO – Standard Error** |
|---|---|---|
| Weibull | *4434.7* | *68.1* |
| Lognormal | *4458.3* | *62.6* |
| Gamma | *4455.3* | *65.6* |

***Supplementary Table 3:*** *Model outputs for the three different distributions used to model the incubation period.*

| **Age Group** | **Distribution** | **Mean Incubation Period** | **Median Incubation Period** | **95th Percentile of the Incubation Period** |
|---|---|---|---|---|
| 3 to 17 years | *Lognormal* | *2.55 (95% CrI: 2.32, 2.83)* | *2.21 (95% CrI: 1.93, 2.52)* | *5.37 (95% CrI: 4.77, 6.08)* |
| 18 to 34 years | *Lognormal* | *2.58 (95% CrI: 2.44, 2.82)* | *2.25 (95% CrI: 2.1, 2.49)* | *5.36 (95% CrI: 4.86, 5.84)* |
| 35 to 44 years | *Lognormal* | *2.57 (95% CrI: 2.42, 2.76)* | *2.23 (95% CrI: 2.08, 2.42)* | *5.34 (95% CrI: 4.89, 5.81)* |
| 45 to 54 years | *Lognormal* | *2.55 (95% CrI: 2.4, 2.69)* | *2.22 (95% CrI: 2.08, 2.36)* | *5.28 (95% CrI: 4.77, 5.68)* |
| 55 to 64 years | *Lognormal* | *2.51 (95% CrI: 2.37, 2.62)* | *2.15 (95% CrI: 2.03, 2.25)* | *5.31 (95% CrI: 4.95, 5.69)* |
| 65 to 74 years | *Lognormal* | *2.59 (95% CrI: 2.48, 2.72)* | *2.23 (95% CrI: 2.13, 2.34)* | *5.48 (95% CrI: 5.17, 5.89)* |
| 75 years and over | *Lognormal* | *2.53 (95% CrI: 2.38, 2.66)* | *2.18 (95% CrI: 2.03, 2.29)* | *5.38 (95% CrI: 5.01, 5.81)* |
| 3 to 17 years | *Gamma* | *2.55 (95% CrI: 2.31, 2.85)* | *2.3 (95% CrI: 2.03, 2.62)* | *5.19 (95% CrI: 4.63, 5.74)* |

| | | | | |
|---|---|---|---|---|
| 18 to 34 years | *Gamma* | *2.57 (95% CrI: 2.42, 2.82)* | *2.33 (95% CrI: 2.16, 2.6)* | *5.2 (95% CrI: 4.76, 5.6)* |
| 35 to 44 years | *Gamma* | *2.56 (95% CrI: 2.41, 2.75)* | *2.32 (95% CrI: 2.16, 2.52)* | *5.15 (95% CrI: 4.71, 5.54)* |
| 45 to 54 years | *Gamma* | *2.54 (95% CrI: 2.39, 2.68)* | *2.31 (95% CrI: 2.15, 2.45)* | *5.08 (95% CrI: 4.62, 5.42)* |
| 55 to 64 years | *Gamma* | *2.5 (95% CrI: 2.37, 2.61)* | *2.25 (95% CrI: 2.12, 2.36)* | *5.15 (95% CrI: 4.86, 5.44)* |
| 65 to 74 years | *Gamma* | *2.59 (95% CrI: 2.48, 2.72)* | *2.32 (95% CrI: 2.21, 2.45)* | *5.34 (95% CrI: 5.08, 5.66)* |
| 75 years and over | *Gamma* | *2.53 (95% CrI: 2.38, 2.66)* | *2.27 (95% CrI: 2.12, 2.41)* | *5.2 (95% CrI: 4.88, 5.54* |
| 3 to 17 years | *Weibull* | *2.55 (95% CrI: 2.25, 2.87)* | *2.34 (95% CrI: 2.04, 2.65)* | *5.25 (95% CrI: 4.44, 6.12)* |
| 18 to 34 years | *Weibull* | *2.57 (95% CrI: 2.39, 2.86)* | *2.37 (95% CrI: 2.18, 2.65)* | *5.31 (95% CrI: 4.83, 5.92)* |
| 35 to 44 years | *Weibull* | *2.54 (95% CrI: 2.35, 2.74)* | *2.35 (95% CrI: 2.16, 2.56)* | *5.12 (95% CrI: 4.65, 5.63)* |
| 45 to 54 years | *Weibull* | *2.5 (95% CrI: 2.31, 2.66)* | *2.32 (95% CrI: 2.11, 2.49)* | *5.03 (95% CrI: 4.6, 5.46)* |
| 55 to 64 years | *Weibull* | *2.5 (95% CrI: 2.36, 2.61)* | *2.27 (95% CrI: 2.13, 2.39)* | *5.26 (95% CrI: 4.96, 5.56)* |
| 65 to 74 years | *Weibull* | *2.62 (95% CrI: 2.5, 2.77)* | *2.38 (95% CrI: 2.25, 2.53)* | *5.54 (95% CrI: 5.25, 5.88)* |
| 75 years and over | *Weibull* | *2.53 (95% CrI: 2.36, 2.68)* | *2.31 (95% CrI: 2.14, 2.47)* | *5.25 (95% CrI: 4.89, 5.64)* |

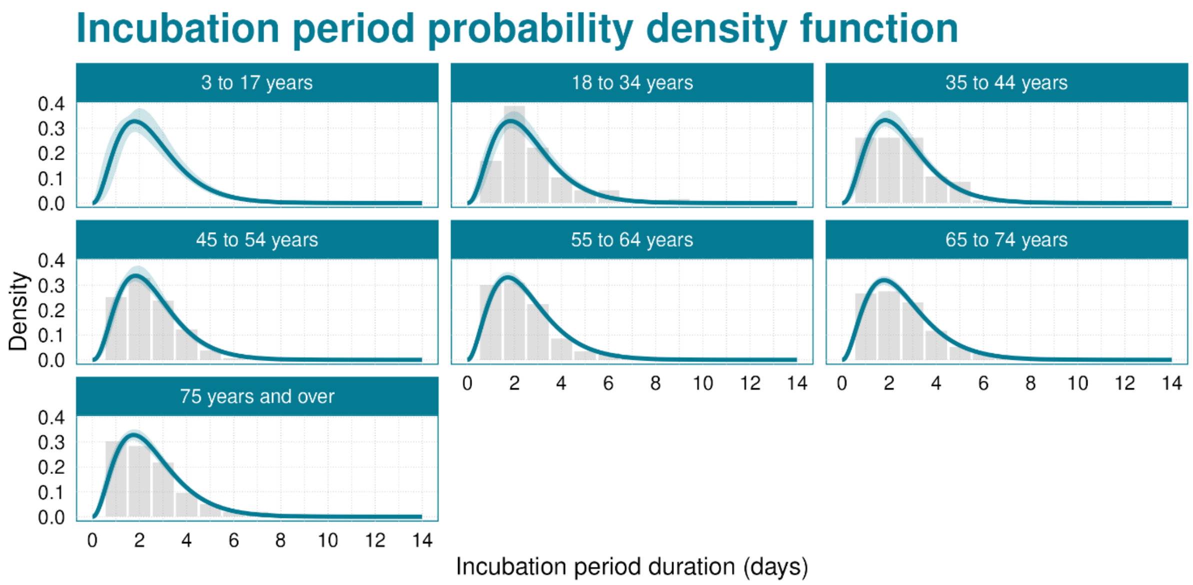


***Supplementary Figure 3:*** *Incubation period distributions estimated using the gamma distribution. The blue line shows the median of the posterior distribution, and the blue shaded region is the 95% credible interval. The grey histogram shows the raw data (the 3 to 17 years age group data are masked due to low counts).*

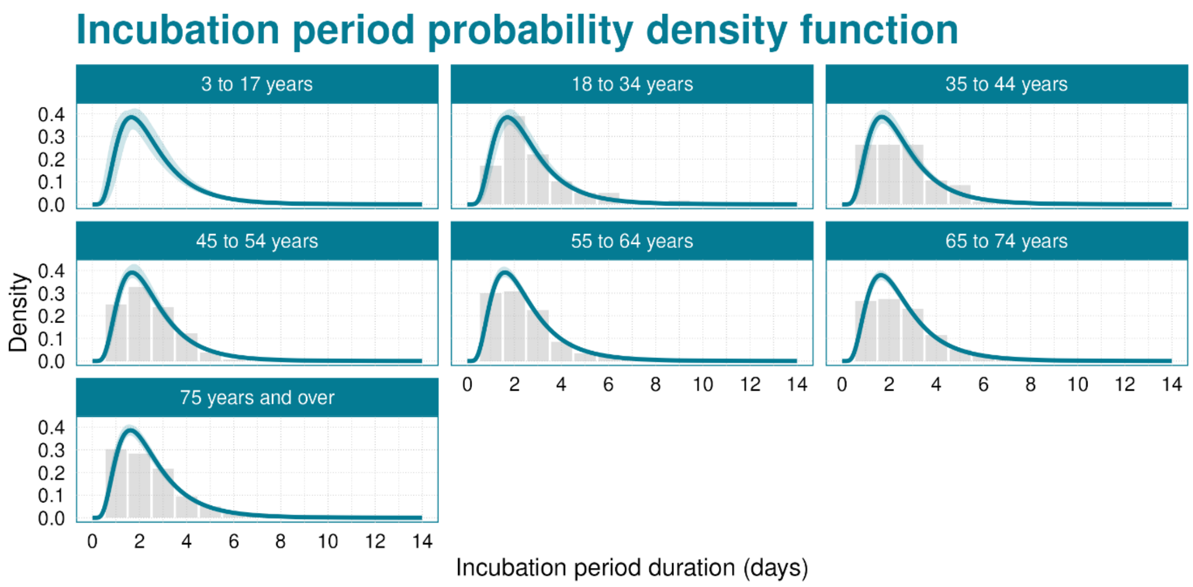


***Supplementary Figure 4:*** *Incubation period distributions estimated using the lognormal distribution. The blue line shows the median of the posterior distribution, and the blue shaded region is the 95% credible interval. The grey histogram shows the raw data (the 3 to 17 years age group data are masked due to low counts).*
